# Assessing the Relationship Between Zero-Dose Communities and Access to Selected Primary Healthcare Services for Children and Pregnant Women in Emergency Settings

**DOI:** 10.1101/2022.08.06.22278500

**Authors:** MP Suprenant, E Nyankesha, R Moreno-Garcia, V Buj, A Yakubu, F Shafique, MH Zaman

## Abstract

In this study the authors examine the relationship between “zero-dose” communities and access to other healthcare services. This was done by first ensuring the first dose of the Diphtheria Tetanus and Pertussis vaccine was a better measure of zero-dose communities than the measles-containing vaccine. Once the best variable was selected, it was used to examine the association with access to primary healthcare services for children and pregnant women residing in the Democratic Republic of Congo, Afghanistan, and Bangladesh, each of which are currently experiencing emergencies of various contexts and degrees. These services were divided into: a) unscheduled healthcare services such as birth assistance as well as seeking care and treatment for diarrheal diseases and cough/fever episodes and b) other scheduled health services such as antenatal care visits and vitamin A supplementation. Using the most recent Demographic Health Survey data from each country (2014: Democratic Republic of Congo, 2015: Afghanistan, 2018: Bangladesh), data was analyzed via Chi Squared analysis or Fischer’s Exact Test. If results were significant, a univariate linear regression analysis was performed to examine if the noted association was linear. While the linear relationship observed between children who had received the first dose of the Diphtheria Tetanus and Pertussis vaccine (the reverse to zero-dose communities) and coverage of other vaccines was expected, the results of the regression analysis depicted an unexpected split in behavior between scheduled (and birth assistance) and unscheduled illness treatment services. For scheduled and birth assistance health services, a linear relationship was generally observed. Meanwhile, for unscheduled services associated with infectious disease treatments, a linear relationship was generally not observed. While in our study it does not appear that first dose of the Diphtheria Tetanus and Pertussis vaccine, the best proxy of zero-dose communities, can be used to predict (at least in a direct linear manner) access to some primary (particularly infectious treatments) healthcare services in emergency/humanitarian settings, it can serve as an indirect measure of health services not associated with the treatment of childhood infections such as antenatal care, skilled birth assistance, and to a lesser degree even vitamin A supplementation.

## Introduction

In the midst of the ongoing COVID-19 pandemic, much focus has been placed on access to vaccines, specifically for this virus, highlighting global inequities in access to healthcare and vaccination. This problem unfortunately is not a novel phenomenon and extends beyond COVID-19 vaccines to even the most basic of childhood vaccination services. These communities where children do not receive the suite of basic vaccinations are known as zero-dose communities and are often operationally defined as locations where children have not received the first dose of a Diphtheria, Tetanus and Pertussis vaccine (DTP1), according to GAVI.^1^

Vaccination against diseases such as Measles, Polio, Tuberculosis and of course DTP is a critical aspect of increasing health protection in early childhood, especially in developing countries where vaccination has accounted for a large decrease in deaths from infectious diseases. As such, studies have looked to elucidate the risk factors associated with children under five not receiving these vaccines and residing in “zero-dose” communities. This has been done in an effort to make better informed predictions about where these zero-dose populations may exist as well as the overall size of these communities so as to better understand how vaccination coverage can be improved so that some of the most vulnerable children can be reached.^2–7^ Despite the number of studies focused on the risk factors associated with zero dose communities as well as strategies to decrease the size of zero dose communities, only a few studies have focused on using zero-dose communities to examine associated impacts on other factors of pediatric healthcare, especially in emergency settings, such as probing the relationship between zero-dose communities and access to both non-infectious and infectious treatment childhood primary healthcare services. Some studies that have attempted to fill this gap and have focused on this relationship provided generalized information for grouped LMICs but did so without any specific focus on emergency settings.^8^ At the same time, Galles et. al have examined the global trend in coverage of various childhood vaccines and reported on association between social demographic index (SDI) and the third dose of DTP (DTP3) noting the sigmoidal relationship between social (SDI) and coverage of this vaccine, while Figueiredo et. al have examined the key factors that correlate to DTP3 coverage between 1980-2010 and 2001-2010 within Africa, the Americas, Europe, Eastern Mediterranean, South-East Asia Region and Western Pacific Region.^9,10^ Furthermore Figueiredo et. al have noted that not only do these socioeconomic factors correlate with DTP3 vaccine coverage but there is also a link between out-of-hospital births and reduced immunization as well as some socioeconomic factors (such as income) relating to utilization of skilled birth assistance. As such, taken together, these studies only transitively draw direct, regional, higher order relationships between zero-dose communities and these correlated health elements through the establishment of a relationship between DTP1 and DTP3 and then DTP3 and other health factors; These metrics all seem to correlate to each other indirectly through socioeconomic factors but no study seems to have directly look at the correlation between DTP1 and treatment for disease, birth related services or vitamin A supplementation.

While these studies provide a helpful analysis, these insights still assume there is already some degree of vaccination access available as these children examined have already received prior doses of DTP. Additionally, as noted before, these results were based on aggregated data from various countries grouped together based on geography or economic level.^8,10^ Although zero-dose communities can exist anywhere, many exist in locations in the midst of humanitarian crisis or armed conflicts which have directly or indirectly resulted in health emergencies.^11^ Given the often geographical or economic inaccessibility of these locations – which is likely one of the drivers of their differential degree of healthcare access even in comparison to nearby/proximal areas – this could lead to them being overlooked especially if the majority of the country or neighboring regions are not in the midst of such a crisis. As such, with this analysis we aim to begin to fill this gap by directly assessing the association between vaccination and access to other healthcare services using data from nationwide surveys for countries representing emergency settings resulting from different conditions. This includes systemic civil unrest and conflict as seen in countries like Afghanistan, more sub-nationally localized civil unrest and conflict as seen in Democratic Republic of Congo (DRC) and localized, migration-based crisis such as the Rohingya crisis in Bangladesh.^12,13^ Here we hypothesize that there is an inverse correlation between zero dose communities and healthcare access and utilization. Furthermore, these findings may provide insight into the possibility of using vaccine uptake as a predictive marker for access to both healthcare services related to non-infectious treatments such as pregnancy and birth related services and healthcare services related to infectious treatments such as treatment for common childhood diseases like cough/fever (indicative of possible respiratory infections) and diarrhea.

## Materials and Methods

Data from DRC, Afghanistan and Bangladesh was collected as part of the Demographic and Health Surveys Program which is publicly available through the program’s website as the most recent complete DHS report at the time of writing. ^14^ Survey responses were organized into topical categories of which the Individual’s Response and Child’s Response categories were specifically examined. These survey responses were collected and reported out at the highest subnational level within each country. To create a nationwide picture of childhood healthcare, we used the combination of all subnational data reports from within a country and classified this as “national” data. In a country like DRC where conflict and resulting humanitarian crisis was more localized, the specific subnational regions undergoing a humanitarian crisis (specifically Kasai-Occidental, Kasai-Oriental, Nord-Kivu and Sud-Kivu) were also separately examined and these locations were classified as “local” data. Data was first collected and analyzed by a Chi Squared test or a Fischer’s Exact Test for sample sizes less than 10 individuals using the GraphPad Prism statistical software using a two-sided test with one degree of freedom for all cases. For significant differences found via the Chi Squared test which were noted as a p value of less than 0.05, survey data was scored and reported as fraction of total individuals who fit each category. This continuous data was then graphed and analyzed in Microsoft Excel to examine the strength of any linear association between variables. Association was determined via a simple univariate linear regression using the R^2^ value to determine the strength of association and an F-statistic (significant at a p value of less than 0.05) to determine if the slope was non-zero..^15^ While we note here our assumption that R squared values of less than 0.2 are considered negligible, 0.2-0.8 weak and 0.8 to 1 as strong, these values are largely arbitrary as various cut off values have been used to demarcate strong vs weak vs negligible degrees of linear associations.^16–18^

### Vaccination Data Calculation

The fraction of children under five who specifically received either the measles vaccine (MCV), DTP1, the third dose of DTP (DTP3), the first dose of the Polio vaccine (Polio1), the third dose of the Polio vaccine (Polio3), or the Bacillus Calmette–Guérin vaccine (BCG), was calculated from the Child’s Response Survey by tallying the number of responses where a child received the vaccine in question according to either a vaccination card (recorded as a 1 or 3) or their caregiver (recorded as a 2) as recorded in the DHS. We took this categorical data and transformed it into the fraction of coverage by dividing the total number of respondents who stated their child was vaccinated by the total number of responses recorded at the provincial administrative level. With the few exceptions of survey responses recorded as an “unknown” or “other” (marked as an 8), this metric is effectively the complementary probability of selecting a child from a zero-dose community out of the region’s population.

This linear regression analysis was performed twice for two different definitions of “vaccinated”. The first, noted as *All Sources*, a child was considered vaccinated if any survey response indicated the child was vaccinated. The second, noted as *Card-only Sources*, a child was considered vaccinated only if the survey indicated they were vaccinated according to their vaccination card such as having the name of the vaccine received on the card and or having the date of vaccination on the card.

### Scheduled Health Services

#### Antenatal Care

As a metric of “Non-Infectious” treatment services, antenatal care in a subnational region was examined. Antenatal care was determined as the fraction of recorded responses where a mother had at least one antenatal care visit divided by the total number of recorded responses to the antenatal care questionnaire.

#### Vitamin A Supplements

As the final example of a non-infectious treatment service, we looked at how vitamin A supplementation varies linearly in comparison to vaccination coverage. To measure the fraction receiving vitamin A supplementation, the fraction of all living children born in the last five years who received vitamin A in the form of an ampoule, a capsule or a syrup in last six months was recorded in the Children’s Response document (noted as a 1). This value was divided by the total number of survey responses in the region to yield the fraction of children under five who had access to vitamin A supplementation in the last six months.

### Unscheduled Health Services

#### Birth Assistance Calculation

Along with antenatal care, birth assistance was also examined though classified as an unscheduled service. Birth assistance was analyzed based on *Skilled Assistance* or the fraction of mothers who responded that they were assisted by a skilled birth attendant (calculated as the fraction of mothers who received assistance during delivery from either a doctor, nurse, health worker or midwife divided by the total number of responses.

#### Access to Care for Diarrhea and Cough/Fever Episodes

Access to treatment was measured as *Access to Medical Care*. This was measured based on the DHS surveys which have direct yes/no responses on if a symptomatic child received medical care which was used to determine this metric (number of responses where a child received medical care divided by total number of respondents who were symptomatic), these surveys also broke down the type/location where treatment was received.

## Results

### Vaccination results

From our analysis, we found that all comparisons made via Chi Squared showed there was a significant association between the coverage pf DTP1 and all other vaccines examined. Additional analysis also determined that there is some linear association between subnational regions vaccine coverage for DTP1 and the coverage of other vaccines in the area, such that as the coverage of DTP1 increases, the coverage of other vaccines also increases. In a single one-to-one comparison between the fraction of children under five who received DTP1 or the fraction of children under five who received the measles containing vaccine (MCV) and either DTP3, the first dose of the Polio vaccine (Polio1), the third dose of the Polio vaccine (Polio3), or the Bacillus Calmette–Guérin vaccine (BCG), this linear association was strongest when DTP1 was used as the independent variable.

#### DRC

More specifically, in the Democratic Republic of Congo, a Chi Squared test was used at the national level to determine if there was a significant association between DTP1 coverage and coverage of DTP3 as well as coverage of MCV, coverage of Polio1, coverage of Polio3, and coverage of BCG. There was a statistically significant association between DTP1 coverage and DTP3 coverage (p< 0.0001), MCV coverage (p <0.0001), Polio 1 coverage (p <0.0001), Polio3 coverage (p < 0.0001) and BCG coverage (p< 0.0001). This same approach was then repeated using MCV instead of DTP with results as follows: There was a statistically significant association between MCV coverage and DTP1 coverage (p <0.0001), DTP3 coverage (p< 0.0001), Polio 1 coverage (p <0.0001), Polio3 coverage (p < 0.0001) and BCG coverage (p< 0.0001). As significant association was observed in all cases, we next looked to see which vaccine proxy DTP1 or MCV produced the better linear association expected between those who are not zero dose children and vaccination status based on the value of R^2^ and checked if the slope of the line is significantly different than 0. For the *All Sources* cases, DTP1 compared to MCV generally produced a better linear fit for DTP3 (p<0.0001, R^2^ = 0.93 vs. p=0.0002, R^2^ =0.80 national; p=0.0310, R^2^ = 0.94 vs. p= 0.0756 R^2^ =0.85 local, respectively) and BCG (p<0.0001, R^2^= 0.98 vs. p<0.0001, R^2^ =0.90 national; p=0.0214, R^2^ = 0.96 vs. p= 0.0051, R^2^ =0.99 local, respectively). For Polio1 and Polio3 both proxy variables performed in a similar fashion (Polio1: p <0.0001, R^2^ = 0.87 for DTP1 vs. p<0.0001, R^2^= 0.87 for MCV at the national level, p=0.0140, R^2^ = 0.97 for DTP1 vs. p=0.0003, R^2^= 1.0 for MCV at the local level) (Polio3: p= 0.1153, R^2^= 0.25 for DTP1 vs. p=0.1213, R^2^= 0.25 for MCV at the national level, p=0.0114, R^2^ = 0.98 for DTP1 vs. p=0.0227, R^2^=0.96 for MCV at the local level). Meanwhile, for the *Card-Only Sources* cases, while both potential proxy variables performed in a fairly similar matter on the national level, on the local level DTP1 produced a stronger association compared to MCV in all cases (DTP3: p= 0.0050, R^2^ = 0.60 vs. p= 0.0070, R^2^ = 0.57 national, p= 0.0887, R^2^ = 0.83 vs. p= 0.1630, R^2^ = 0.70 local, respectively; BCG: (p= 0.0049, R^2^= 0.60 vs. p=0.0076, R^2^ = 0.57 national, p= 0.0753, R^2^ = 0.86 vs. p=0.1429, R^2^ = 0.73, local, respectively; Polio1: p= 0.0051, R^2^ = 0.60 vs. p= 0.0081, R^2^ = 0.56 national, p= 0.0789, R^2^ = 0.85 vs. p= 0.1478, R^2^ = 0.73 local; and Polio3: p= 0.0058, R^2^= 0.59 vs. p= 0.0078, R^2^ = 0.56 national, p=0.0882, R^2^ = 0.83 vs. p=0.1624, R^2^ = 0.70 local. Thus, it was observed for nearly all cases that the linear association was both stronger and with a slope more likely to be significantly different than zero for all vaccines examined when including caregivers’ responses in the estimation of vaccination coverage (*All Sources* data) compared against the relationship observed when only using coverage information if it came from a vaccine card (*Card-Only Sources* data). The notable exception to this is in the case of Polio3 where at the national level the card only data produced a fit with an R^2^ value nearly double of the *All sources* data. Furthermore, it was observed that the relationship was at least as strong and often stronger in the specific subnational regions where conflict is more apparent compared to the relationship seen throughout all subnational regions in the country. [Figure 1] This trend was also observed with MCV. [Figure S1]

**Figure 1.**
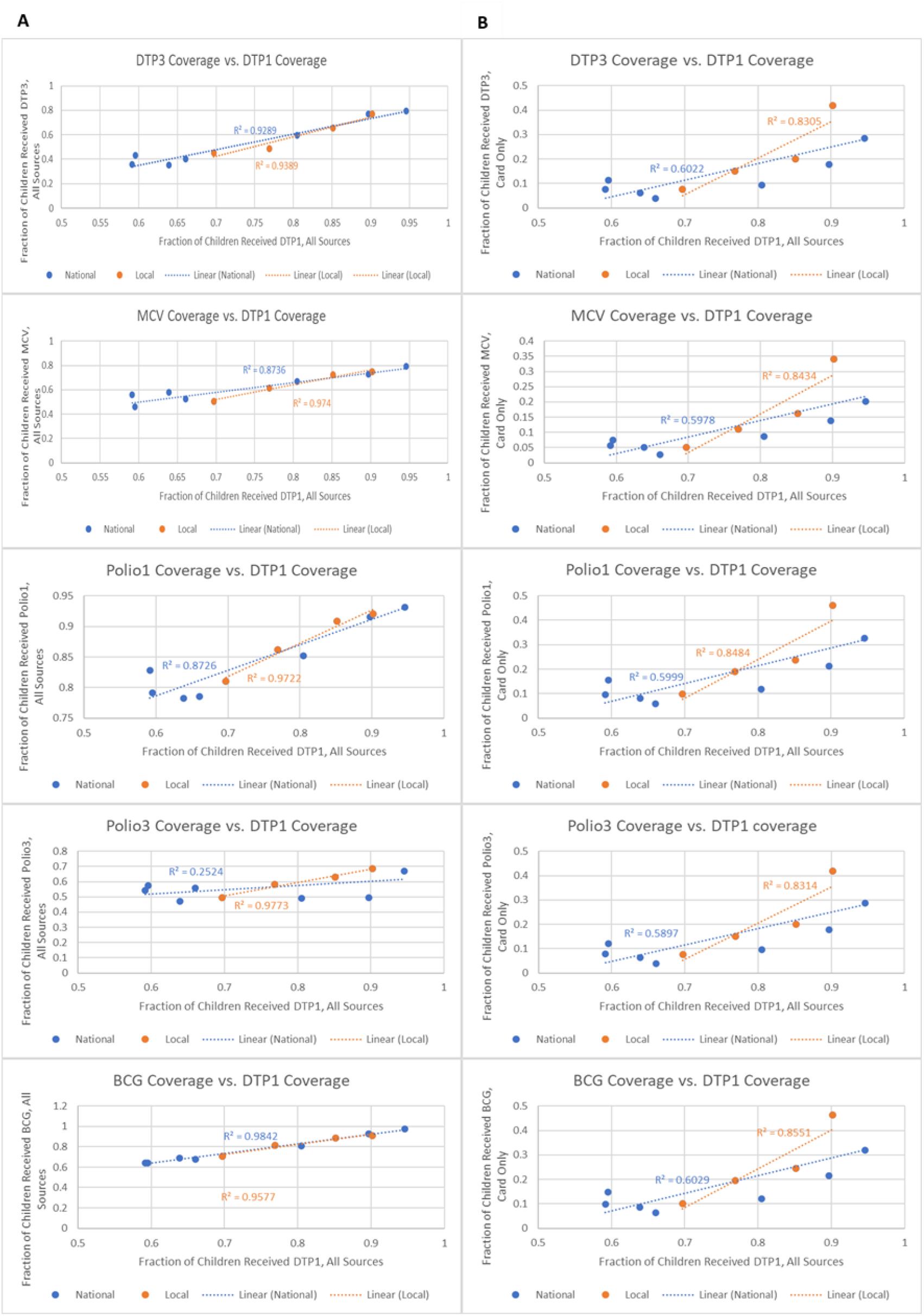
Univariate linear regression between DTP1 vaccination coverage and coverage for other common vaccines in DRC. Column A shows how the inclusion of caregivers’ responses improve the linear fit compared to the relationships in column B which only considered a child vaccinated if their vaccination card contained the appropriate vaccine information. In this case, a difference was observed between the strength of the relationship observed in regions with conflict compared to the entire country.

#### Afghanistan

In Afghanistan, Chi squared results were as stated: There was a statistically significant association between DTP1 coverage and DTP3 coverage (p< 0.0001), MCV coverage (p <0.0001), Polio 1 coverage (p <0.0001), Polio3 coverage (p < 0.0001) and BCG coverage (p< 0.0001). The results were similar for MCV as there was a significant association between MCV coverage and DTP1 coverage (p <0.0001), DTP3 coverage (p< 0.0001), Polio 1 coverage (p <0.0001), Polio3 coverage (p < 0.0001) and BCG coverage (p< 0.0001). Comparing the linear fits of all sources data, DTP1 resulted in a better linear fit for DTP3 coverage (DTP1 p<0.0001, R^2^ = 0.91 vs. MCV: p<0.0001, R^2^ = 0.82) and BCG coverage (DTP1: p<0.0001, R^2^ = 0.95 vs. MCV: p< 0.0001 R^2^ = 0.78). Conversely, MCV resulted in a stronger fit for both Polio1 (DTP1: p<0.0001, R^2^ = 0.50 vs. MCV: p<0.0001 R^2^ = 0.64) and Polio3 coverage (DTP1: p< 0.0001, R^2^ = 0.56 vs. MCV: p<0.0001 R^2^ = 0.66). For Card-Only sources the results were as follows: DTP3: p<0.0001, R^2^ =0.61 for DTP1; p <0.0001, R^2^ = 0.50 for MCV; BCG: p<0.0001, R^2^ =0.64 for DTP1; p<0.0001, R^2^ = 0.48 for MCV; Polio1: p<0.0001, R^2^ =0.63 for DTP1; p<0.0001, R^2^ =0.48 for MCV; Polio3: p<0.0001, R^2^ =0.61 for DTP1; p<0.0001, R^2^ =0.49 for MCV showing the improved fit when DTP1 was used compared to MCV. Again, we generally observed a stronger linear association with DTP1 when examining the *all sources* data compared against the *card-only sources* data with the exception of both doses of the Polio vaccine. [Figure 2] This trend was also maintained when MCV was used as the independent variable [Figure S2]

**Figure 2.**
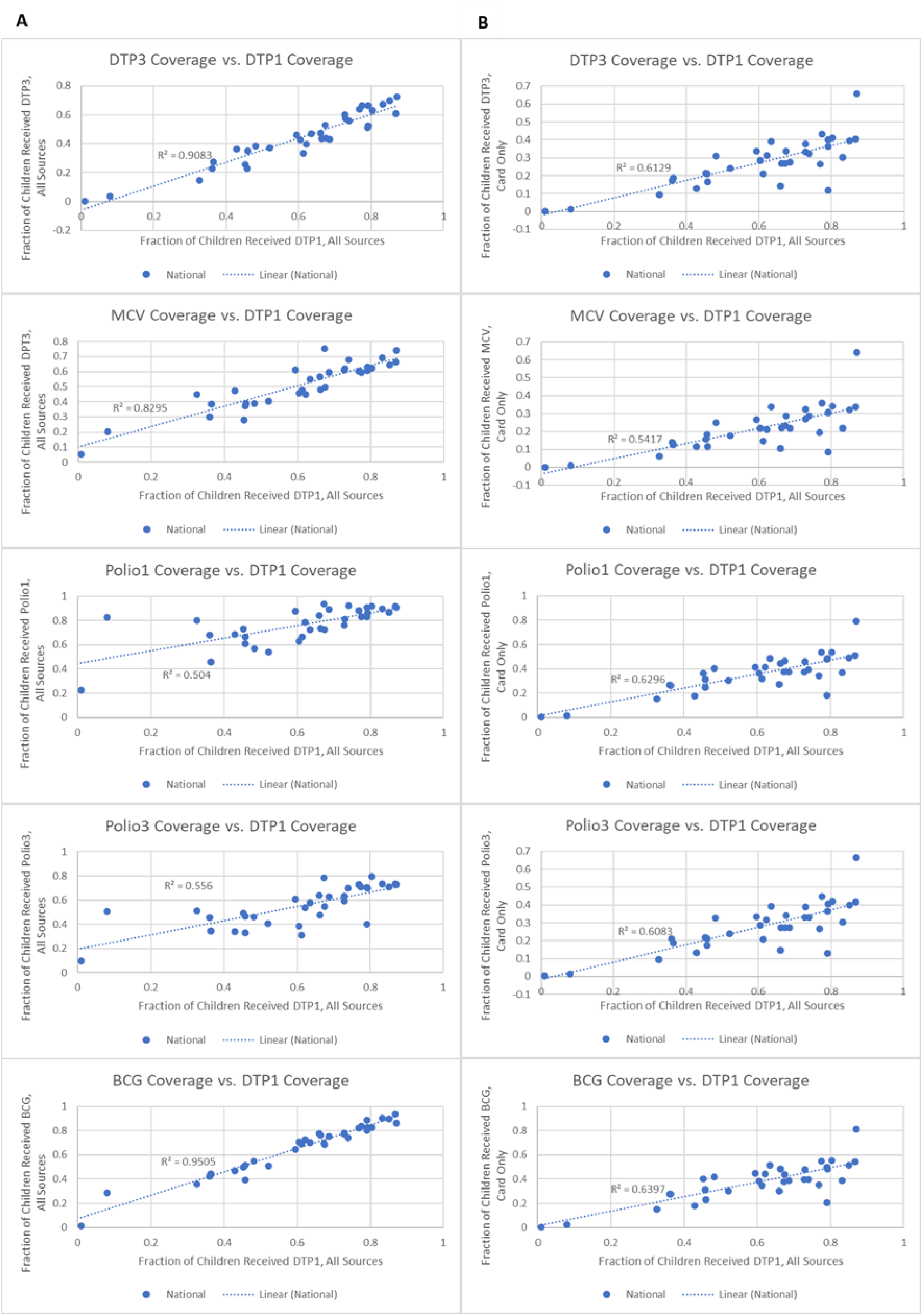
Univariate linear regression between DTP1 coverage and coverage for other common vaccines in Afghanistan. Column A shows how the inclusion of mothers’ responses improve the linear fit compared to the relationships in column B which only considered a child vaccinated if their vaccination card contained the appropriate vaccine information. This trend was not observed for either dose of the Polio vaccine, though the strengths of the fit were similar for both metrics used.

#### Bangladesh

Lastly, in Bangladesh the following results were observed. There was a statistically significant association between DTP1 coverage and DTP3 coverage (p< 0.0001), MCV coverage (p <0.0001), Polio 1 coverage (p <0.0001), Polio3 coverage (p < 0.0001) and BCG coverage (p< 0.0001). The results were similar for MCV as there was a significant association between MCV coverage and DTP1 coverage (p <0.0001), DTP3 coverage (Fisher’s exact test, p< 0.0001), Polio 1 coverage (Fisher’s exact test, p <0.0001), Polio3 coverage (Fisher’s exact test, p < 0.0001) and BCG coverage (Fisher’s exact test, p< 0.0001).Here, DTP1 resulted in a stronger linear association among the *All Sources* data compared to MCV for DTP3 coverage (p=0.0024, R^2^ = 0.81 vs. p = 0.0463, R^2^ =0.51, respectively), Polio1 coverage (p<0.0001, R^2^ = 0.98 vs. p=0.0718, R^2^ =0.44, respectively), Polio3 coverage (p=0.0040, R^2^ = 0.77 vs. p=0.0267, R^2^ =0.59, respectively) and BCG coverage (p= 0.0019, R^2^ = 0.82 vs. p= 0.3152, R^2^ =0.17, respectively).

For the Card Only sources results were similar with DTP1 performing better than MCV for DTP3 coverage (p=0.2838, R^2^ = 0.19 vs. p = 0.2406, R^2^ =0.22, respectively), Polio1 coverage (p=0.1897, R^2^ = 0.27 vs. p=0.4810, R^2^ =0.09, respectively), Polio3 coverage (p=0.2950, R^2^ = 0.18 vs. p=0.2677, R^2^ =0.20, respectively) and BCG coverage (p= 0.2321, R^2^ = 0.23 vs. p= 0.6343, R^2^ =0.04, respectively). As with the survey results from both DRC and Afghanistan, a stronger linear association was observed when examining the *all sources* data compared against the *card-only sources* data when either MCV was used as the independent variable [Figure S3] or DTP1 was used as the independent variable [Figure 3] though even with DTP1 as the independent variable for the Card-Only sources case regardless of R^2^ values, no slope was significant different than 0 though in general the use of DTP1 resulted in all R^2^ values being increased. Especially noticeable in Figure 3 were the individual relationships between DTP1 and DTP3, MCV, and Polio3. For these three vaccines, the R^2^ values were below the cut off for a weak linear relationship to be observed when only vaccination card data was considered, but after the inclusion of responses from mothers all displayed linear relationships.

**Figure 3.**
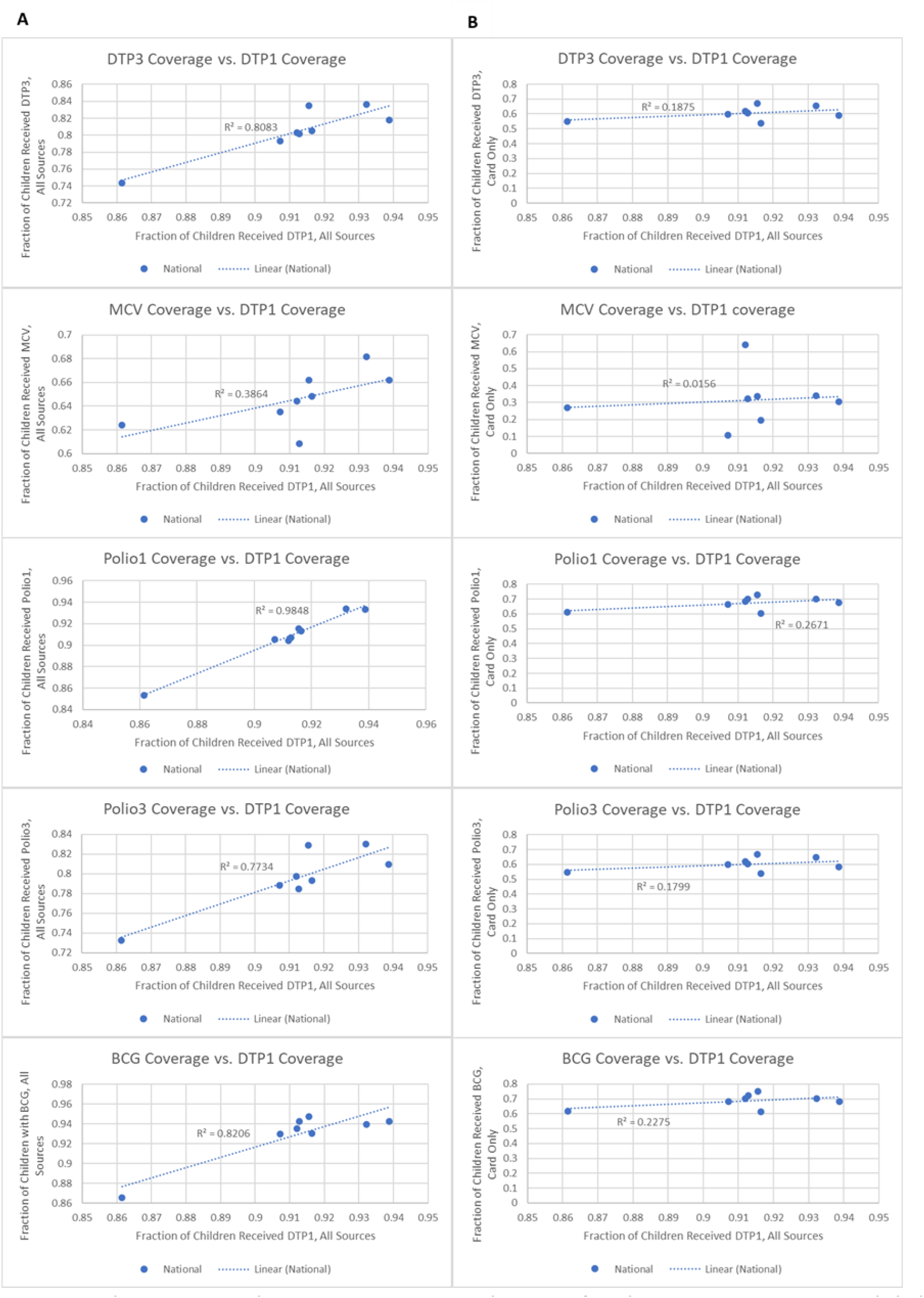
Univariate linear regression between DTP1 coverage and coverage for other common vaccines in Bangladesh. Column A shows how the inclusion of mothers’ responses improve the linear fit compared to the relationships in column B which only viewed a child as vaccinated if their vaccination card contained the appropriate information. This trend was especially apparent for DTP3, MCV, and Polio3 as the *card only* metric led to no linear relationship being observed, while the *all sources* data saw linear relationships for all cases.

### Treatment for Diarrhea and Cough/fever

#### DRC

In the DRC, a Chi Squared analysis showed there was a significant association between DTP1 and medical treatment for diarrhea at the national level (p= 0.0006) and the specific local regions examined (p = 0.0008). Despite both having significant associations, there was a noticeable difference between the observed relationship with diarrhea treatment when examining the general medical treatment data from facilities and community health workers throughout the country and the relationship when observed only in the local, specific emergency regions in the country. In these specific subnational areas, a strong linear relationship was seen between the fraction of children under five who received DTP1 and general medical treatment for diarrhea (p=0.0212, R^2^=0.96). When examined over the whole country however, no linear relationships were observed for diarrhea treatment (p=0.6901, R^2^=0.02) indicating a split in behavior potentially due to sample size.

For an examination of cough/fever treatment, the chi squared analysis again showed a significant association at the national level (p <0.0001) but not a significant association at the local level (p=0.0710). In these cases, no linear relationship between cough/fever treatment and DTP1 coverage was observed in either the “local” data set or the national data set (p = 0.9857, R^2^=0.00 for the national case; p =0.9376, R^2^=0.00 for the local case).

#### Afghanistan and Bangladesh

Trends in Afghanistan and Bangladesh, however behaved more akin to the DRC national level data. For Afghanistan, the Chi Squared analysis showed a significant association between DTP1 and medical treatment from facilities and community health workers for diarrhea (p <0.0001) and medical treatment from facilities and community health workers for cough/fever (p <0.0001) while for Bangladesh no significant association was seen between DTP1 and medical treatment for diarrhea (p= 0.2923) nor for cough/fever medical treatment (p = 0.3781). While there was an association for Afghanistan, further examination revealed it was not a linear association for diarrhea (Afghanistan p=0.2360, R^2^=0.04) nor for cough/fever (Afghanistan p=0.4708, R^2^=0.02) due to low R^2^ values and the slope of the relationship not being significantly different than zero. The lack of linear association for both cases in Bangladesh (diarrhea p=0.7668, R^2^ =0.02 cough/fever p=0.8648, R^2^ =0.01) was expected given the lack of any significant association expected via the Chi Squared results. [Figure 4]

**Figure 4.**
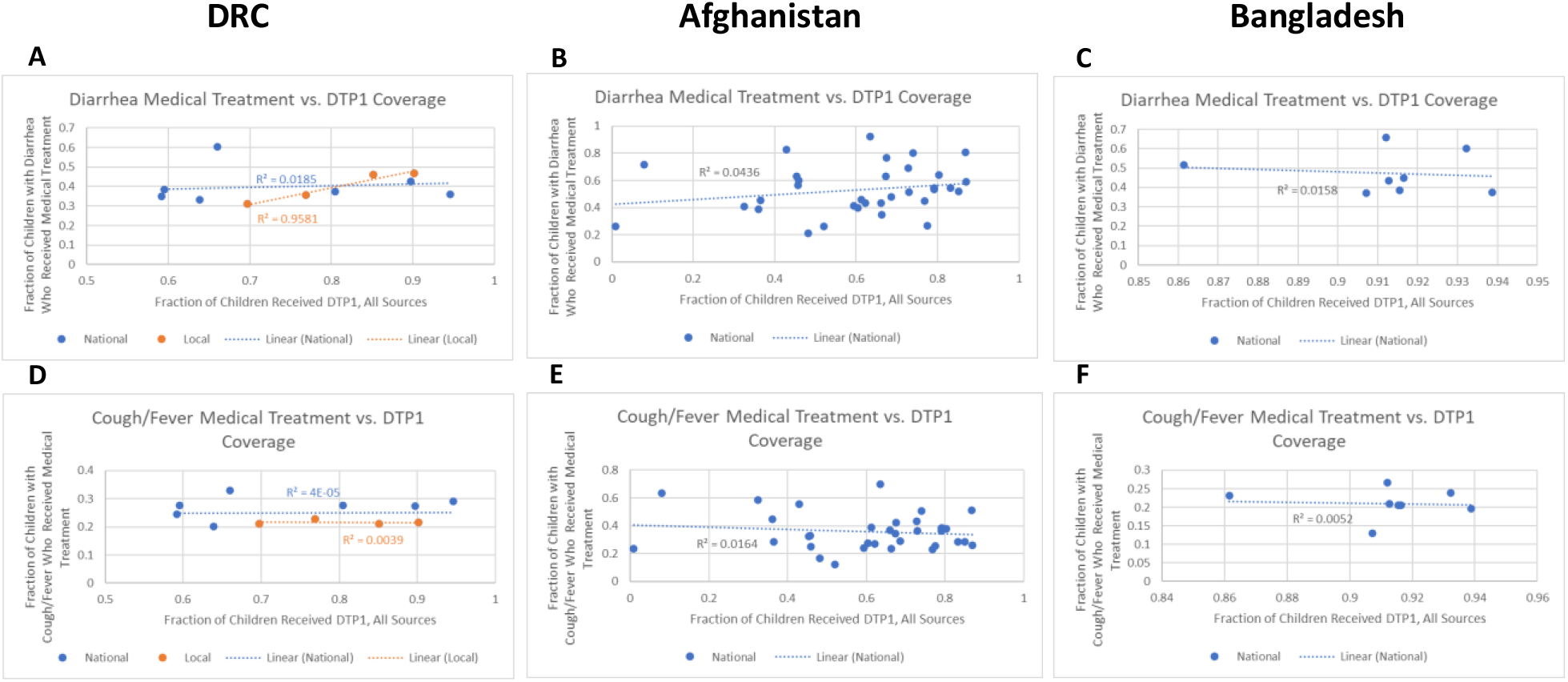
Univariate linear regression between children under five who received DTP1 and medical treatment for diarrhea and for cough/fever. **Columns show results for** DRC (A, D), Afghanistan (B, E) and Bangladesh (C, F), respectively. When examined at the subnational level throughout the whole country no linear trends were observed. However, specific crisis regions within DRC did show a strong linear relationship between DTP1 and medical treatment for diarrhea.

### Antenatal Care

The results for antenatal care followed the same trend seen for vaccination coverage: as the fraction of children covered by the vaccine in question increased within each subnational region, the fraction of mothers who received antenatal care visits also increased. [Figure 5]

**Figure 5.**
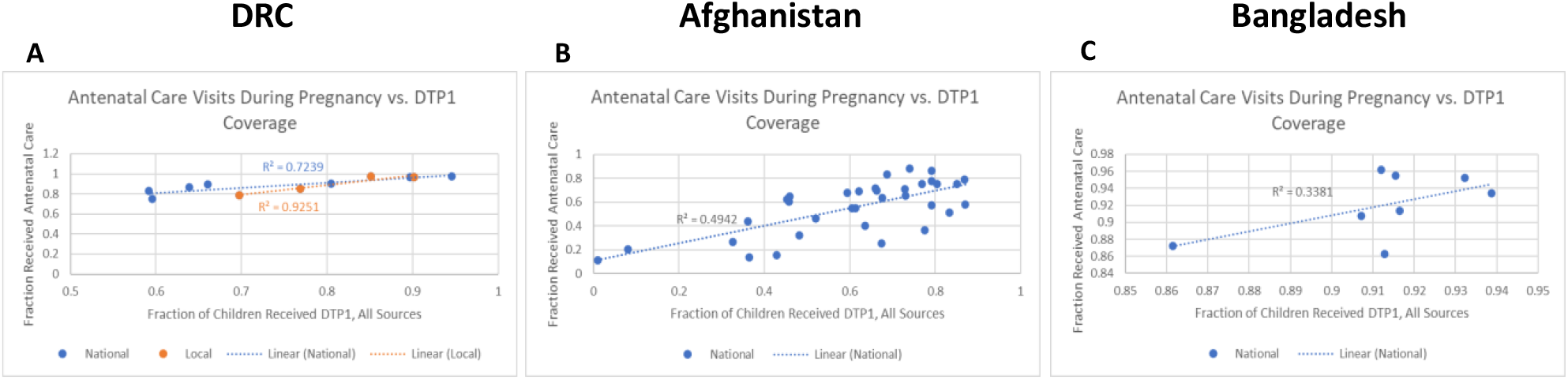
Comparison of degree of linear association observed between DTP1 coverage and women who received antenatal care. Local data points in DRC (A) refer to subnational regions in DRC where emergency conditions are more acute, specifically Kasai-Occidental, Kasai-oriental, Nord-kivu and Sud-kivu, while the national set encompasses all subnational data points. The degree of linear association was strongest in these specific regions in the DRC compared to the country as a whole (R^2^ =0.93 vs. R^2^ =0.72). Meanwhile Afghanistan (B) and Bangladesh (C) still saw weak linear associations between DTP1 coverage and antenatal care visits (R^2^ =0.49, and R^2^ =0.34 respectively).

#### DRC

Chi Squared analysis for antenatal care showed a significant association between DTP1 and antenatal care nationally (p<0.0001) and sub nationally (p<0.0001). Additional examination into the association showed that in the DRC, an almost perfect positive linear association was observed between increasing DTP1 vaccination coverage and increasing the fraction of women who received antenatal care visits within the specific subnational regions in DRC (p=0.0382, R^2^=0.93). When examined throughout all subnational areas in the national DRC dataset, a linear trend with a slope significantly different than zero was still observed (p= 0.0009, R^2^=0.72).

#### Afghanistan

Likewise in Afghanistan the results from the Chi Squared analysis showed a similarly significant association between DTP1 coverage and antenatal care (p < 0.0001). Despite the similar results in general association to those observed in the DRC, additional analysis for Afghanistan did not produce as strong of a linear trend comparatively, though a linear association with a non-zero slope was still observed (p<0.0001, R^2^=0.49).

#### Bangladesh

Lastly, antenatal care results from Bangladesh again showed a significant (albeit less so) association between DTP1 coverage and antenatal care (p = 0.0012). However, unlike both the DRC and Afghanistan data sets, while the line of best fit has an R^2^ value of 0.34 indicating at least a weak linear association it did not have a slope that was significantly different than zero (p=0.1306).

### Birth Assistance

To further examine the potential relationship between unscheduled services and DTP1 vaccination status, birth assistance was also examined with findings similar to those for antenatal care as shown in Figure 6.

**Figure 6.**
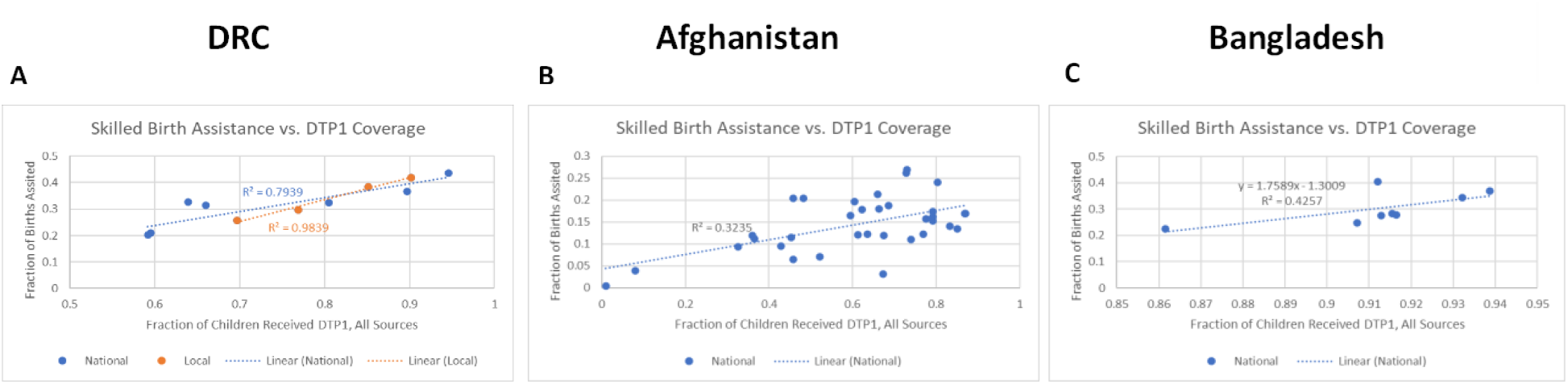
Linear regression between DTP1 and birth assistance in DRC (A), Afghanistan (B), and Bangladesh (C). Skilled birth assistance was considered as receiving assistance from a doctor, nurse, midwife, or healthcare worker. Linear associations were observed in DRC (A), Afghanistan (B) and Bangladesh (C) for skilled birth assistance (R^2^ =0.79, 0.32, and 0.43 respectively).

#### DRC

In the data from the DRC, the Chi Squared test returned a significant association at the national level between DTP1 and receiving birth assistance from a skilled medical worker (p<0.001) as well as at the local level (p<0.001).

#### Afghanistan

In Afghanistan Chi Squared results were analogous to those in the DRC with significant association observed between DTP1 coverage and birth assistance from a skilled medical worker (p<0.001).

#### Bangladesh

The association in Bangladesh between DTP1 coverage and skilled birth assistance was significant (p = 0.0141). Despite this less significant association, a linear association was still observed for birth assistance from a skilled medical provider though the slope of the line was not significantly different than zero (p=0.0795, R^2^ =0.43).

### Vitamin A Supplementation

To see if the relationship observed between DTP1 and non-infectious treatment services held for those services beyond birth and pregnancy related ones, vitamin A supplementation was also examined with results shown in Figure 7.

**Figure 7.**
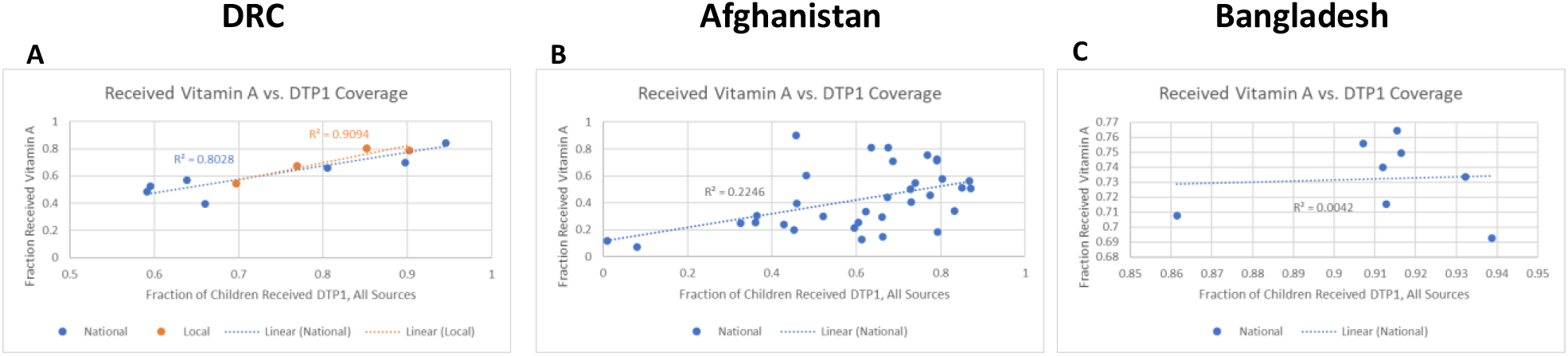
Comparison of degree of linear association observed between DTP1 coverage and vitamin A supplementation. (A), Afghanistan (B) and Bangladesh (C). This association was seen in the local and national data sets in DRC (A). In Afghanistan (B), the association was weaker, producing an R^2^ indicating a weak association (R^2^ =0.22). However, the data set for Bangladesh (C) showed no linear association, appearing to be nearly perfectly separate, dissociated variables (R^2^ = 0.00).

#### DRC

In the DRC as with the prior variables examined, there was a significant association between DTP1 and Vitamin A supplementation via Chi Squared test at the national level (p <0.0001) and local level (p<0.0001). Further examination of the data and the association yielded a strong linear trend when DTP1 was used and compared against the fraction of children who received vitamin A in the last six months. This association was strongest when restricted to the specific regions where conflict was most present, producing an R^2^ value of 0.90 with a p = 0.0464 compared to an R^2^ of 0.80 with p= 0.0002 seen at the national level.

#### Afghanistan

In Afghanistan, the initial Chi Squared results also indicated a significant association between DTP1 and Vitamin A supplementation (p<0.0001) and the fraction who received Vitamin A supplements in the last six months also demonstrated the association was a weak linear association (R^2^=0.22, p=0.0046) with the fraction who received DTP1. However, unlike in DRC, the R^2^ value was closer to the cutoff between no linear association and a weak linear association, showing the strength was much weaker in this data set and might ultimately be a more complicated relationship.

#### Bangladesh

Lastly, in Bangladesh, despite a significant association recorded according to the Chi Squared analysis between DTP1 and Vitamin A supplementation (p<0.0001), the association did not appear to be a linear as it produced an R^2^ =0.00 with p= 0.8792. In this case, the two variables were nearly perfectly linearly dissociated.

## Discussion

### Vaccination coverage

While DTP1 vaccine coverage showed a similar degree of associations (both generally via Chi Squared and linearly via linear regression) to childhood coverage of DTP3, MCV, BCG, Polio1 and Polio3 vaccines in DRC and Afghanistan as MCV vaccine coverage showed, more linear associations were observed in Bangladesh when DTP1 was examined instead of MCV. While this was expected given the operational definition of zero-dose communities, this confirmation combined with the operation definition led to the selection of DTP1 as the main variable for analysis in our work going forward instead of other vaccines such as MCV. ^1^ In this case it was expected that as DTP1 coverage increased (and therefore as Zero-dose populations decreased) the coverage of routine vaccination would also increase in all three countries. Of particular interest is the breakdown between how vaccination status was assessed, namely either based solely on a vaccine card (which has been notated as *Card-only Sources*) or supplemented with insight from the mother of the child in question (notated as *All Sources*). While vaccination card data is the standard approach for assessing if a child has actually received a specific vaccine eliminating the need for a mother to accurately recall which vaccine or which dose of a vaccine her child may have received, we found there was a stronger association and better ability to estimate vaccination coverage from DTP1 coverage if the mother’s response was taken into consideration, as in all three countries examined, this *All Sources* metric resulted in an improved linear fit via a larger R^2^ value. While potentially less accurate in the “actual” vaccination coverage values, this approach within each subnational region still has its benefits for planners as the *All Sources* metric can act as a sort of “ideal” coverage value where it can be assumed this value represents the best possible vaccination case if every mother’s response and memory recall was completely accurate. As such, this provides a useful metric for what proportion of the population in the area is not actively vaccinated (the remaining population) which can help health care workers and planners better prioritize which regions are more in need of vaccination assistance for a range of vaccines in a timely manner based simply on the coverage of one dose of a single vaccine. While this metric might not provide concrete estimates of vaccination coverage for academic or research purposes, we nonetheless believe such a metric might still be of practical use for fieldworkers on the ground.

### Birth Assistance

As the goal for improving healthcare access would be for women to receive assistance from a healthcare provider rather than anyone, we examined the relationship between DTP1 coverage and skilled birth assistance. Here, we found that a positive linear relationship was observed, a result largely expected due to prior observations between the correlation of DTP3 and births attended by health staff and is supported additionally by our findings that antenatal care also was linearly related to DTP1 coverage which would likely produce similar results.^10^ This seems to indicate that similar types of treatment services will behave in a similar manner to each other even if they are grouped dissimilarly in terms of their more arbitrary timing/schedule classification. That said, explanations behind this relationship are partially complicated by the fact that in addition to socio-economic status and geographical factors there are also a lot of social and behavioral factors which determine whether a woman will seek skilled birth care including education level of her mother-in-law and any “taboos” around birth and pregnancy common in the area. Thus, based on the observations made on this data set, utilization of skilled birth assistance within a subnational region of a country would be able to be estimated by knowing the population size of zero-dose children who reside in this portion of the country.

### Unscheduled Services vs. Scheduled Services

With the selection of our metric for a zero-dose community analogue variable determined and confirmed to match prior approaches, we began to examine if there was any relationship between DTP1 coverage and utilization/access to other health care services. Various types of services were considered and split into two different overarching groups, namely scheduled services, and unscheduled services. We defined unscheduled services as medical services needed for sudden treatments such as illness-based services like those for the treatment of diseases, such as the treatment of common childhood illnesses such as diarrhea or possible respiratory infections (noted in the data as cough/fever) as well as non-illness-based services such as assistance during birth. Conversely, scheduled services were defined as medical services that could be planned for such as antenatal care and nutritional supplementation services like those for vitamin A. With this distinction in place, a split in behavior was generally able to be observed between how zero-dose communities relate to access to care for other services. Based on the data sets used in this analysis and the geographical resolution provided, we noticed that there did not seem to be any linear relationship between these communities and access to care for unscheduled services related to illness, while there was an observed linear relationship when non-infectious/scheduled treatment services were examined. While at least a weak linear relationship was observed in two of the three countries examined (notably those that are experiencing a crisis with armed conflict as a key component as opposed to a migration-based crisis as is the case in Bangladesh), the R^2^ value was generally lower, indicating that the association was not as prevalent compared to the birth and pregnancy related services observed. That said in all three countries, there always was a significant association between DTP1 coverage and scheduled services even if the association was not linear. This seemed to indicate that not only do zero-dose communities have a different relationship between scheduled and unscheduled services, but these relationships are even more nuanced allowing the linear association to appear more strongly (or even at all) for these pregnancy related services in each group compared to other types of services in each classification, especially in conflict regions. For healthcare planners, this means that by knowing the proportion of children in zero-dose communities within a subnational region of a country, estimates can be made for the degree of access to or utilization of these scheduled services (and birth assistance), with an increasingly more accurate prediction made for antenatal care and skilled birth assistance compared to other services, especially in countries where the crisis is largely conflict driven and more systemic. Furthermore, based on this analysis, it appears that even if the exact size of the zero-dose population is unknown for certain locations, healthcare planners can still work to determine which areas are most in need of assistance in increasing the access to care for scheduled services by determining which locations are likely to have the greatest zero-dose population. This relative, priority order type approach can provide quick insight into the ground situation even in locations where data may be more limited, similar to the “ideal” vaccination approach mentioned earlier in the discussion section. This approach would, however, be insufficient for unscheduled services related to illness treatment due to the lack of observed linearity between zero-dose communities and access to these types of care services based on the data used for this analysis which included both facility and community-based health workers. We note that while a linear relationship was not seen between zero dose communities and infectious treatment services, for the three samples that did result in significant association by Chi Squared analysis (but not linear regression) this could mean that the relationship at this level is more complicated than a linear relationship or simply that such a linear relationship may only be observable at a more local level. As such, future work can further search for a relationship between zero-dose communities (and by proxy vaccination status) and access to infectious treatment healthcare services, especially at an even more local, perhaps district, level.

### Limitations

We note that this analysis does have limitations. One such limitation is the DRC specific data. As some of the emergency conditions are more localized to a few specific subregions in the country (the local dataset) the regression analysis on this portion of the data only had four data points. This means each point contributes more heavily to the line of best fit, making it more susceptible to the impact from any outliers. Furthermore, such a small sample set here means that linear relationships (and any statistical findings) could be observed by chance. To help counter these limitations, analysis was done not only within these few specified local regions but also across all areas of the country at this same government level as a means of comparing this smaller sample size against a more statistically robust data set. In many cases the trends seen in the local data set are similar to the national level, but in a few cases (such as the treatment for diarrhea) there is a difference between these two data sets with the local set resulting in an R^2^ that is ∼50 times larger, most likely indicating that this behavior is due to the small sample size.

Likewise, we note that in this analysis, even the most statistically robust data set (Afghanistan) had only about 20 data points for the linear analysis. While the data sets could be pooled together to create a supranational data set to increase statistical power, we believe this approach would have added additional complications and limitations. One such example is that since the data was collected within each country separately, combining these data points could possibly result in the addition of country-country confounding factors as well as temporal confounding factors given that although each survey was the most recent version, they were each performed over different timeframes. Thus, we opted to keep each country separate and use all the information provided at this level to make the findings as robust as possible despite the ultimately smaller size.

An additional data-based limitation presents itself during the infectious treatment services analysis, specifically with regards to treatment of cough/fever. We note that conjoining cough and fever may result in a flawed analysis since “fever” is usually the first symptom for seeking care, especially in the DRC where malaria is such a high burden. In these locations with a large number of malaria cases, cough would be a secondary symptom, which in addition to rapid diagnostic tests would help distinguish between upper respiratory infections and malaria. This cough/fever metric was used as while the DHS data set had information on fever episodes, there was no information specifically regarding treatment for fever or treatment for cough; treatment data was only recorded as “treatment for cough/fever”. Based on the region and overlap with Malaria, and the lack of a respiratory infection section, this most likely is done specifically to filter out treatment for Malaria and focus just on potential respiratory infections. Despite this assumption, in an effort to be more direct and clearer in our analysis the name was left unchanged from the titles in the DHS data sets.

Furthermore, while we initial perform a Chi Squared analysis to determine if there is any association between the variables we examined, we note that if there is an association this analysis only looks to see if that association is a simple linear relationship between the variables described. Thus, while linear predictions might not be possible due to a lack of a linear association, that does not necessarily mean that such an analysis using a different regression approach would result in a lack of association; a different relationship may exist that relates the dependent and independent variables examined in this report, which is what would be expected for the comparisons that saw an association via Chi Squared but not linear regression.

### Conclusion

In this analysis, we analyzed data sets from countries experiencing different types of emergency conditions such as DRC, Afghanistan, and Bangladesh to examine the best proxy for zero dose communities as well as the association between this variable and access to and utilization of other health services, such as those for treating common childhood diseases like diarrhea and cough/fever, which is usually indicative of respiratory infections, antenatal care, birth assistance and vitamin A supplementation. While other papers have examined what factors are related to zero-dose communities and therefore how to best predict where these communities are largest, fewer studies have focused on the relationship between zero-dose communities and its ability to estimate access to other health services in specific emergency settings, a gap we attempted to begin to bridge with this analysis. To this end, we have observed at the subnational level that DTP1 vaccination status was a good indicator for zero-dose communities and resulted in a stronger set of linear relationships compared to MCV for the prediction of coverage for other vaccines. Furthermore, we noticed a split in behaviors depending on the type of services examined. DTP1 coverage saw no linear relationship with unscheduled infectious treatment services and thus could not be used to make predictions on this facet of health, however linear relationships were observed when examining all scheduled health services as well as birth assistance, a non-infectious unscheduled service. Thus, in emergency/humanitarian settings, DTP1 coverage is the best proxy for labeling a community as zero-dose and is linearly associated with access and utilization of antennal care, skilled birth assistance, and to a lesser degree vitamin A supplementation. As such DTP1 coverage can be used as an indirect measure of these metrics, even though this was not the case for the treatment for childhood diseases such as diarrhea and cough/fever which saw no linear association with DTP1 coverage.

## Supporting information

Supplemental Vaccination Results

## Data Availability

All data results are present in the contained manuscript and all data used for the production of said results are available online through the Demographic Health Surveys Program

https://dhsprogram.com/data/

## Acknowledgements

The authors would like to thank Oya Zeren Afsar, Ibrahim Dadari, and Sarah Tougher for their feedback during the revision process of this manuscript.

We would also like to thank Jacqueline Hicks, Ph.D. for her guidance and review of the statistical approaches used in the development of this manuscript.

## References

1. Gavi. Zero-Dose Analysis Card. https://lnct.global/wp-content/uploads/2021/08/Gavi_Zero-dose_AnalysisCard.pdf (2019).

2. Greenwood, B. The contribution of vaccination to global health: Past, present and future. Philos. Trans. R. Soc. B Biol. Sci. 369, (2014).

3. GAVI. Data for Zero-dose: what was done? What did we learn? What next? (2021).

4. Johri, M., Rajpal, S. & Subramanian, S. V. Progress in reaching unvaccinated (zero-dose) children in India, 1992–2016: a multilevel, geospatial analysis of repeated cross-sectional surveys. Lancet Glob. Heal. 9, e1697–e1706 (2021).

5. Deshpande, A. et al. Mapping geographical inequalities in access to drinking water and sanitation facilities in low-income and middle-income countries, 2000–17. Lancet Glob. Heal. 8, e1162– e1185 (2020).

6. Arambepola, R. et al. Using geospatial models to map zero-dose children: Factors associated with zero-dose vaccination status before and after a mass measles and rubella vaccination campaign in Southern province, Zambia. BMJ Glob. Heal. 6, (2021).

7. Wahl, B. et al. Change in full immunization inequalities in Indian children 12-23 months: an analysis of household survey data. doi:10.1186/s12889-021-10849-y.

8. Santos, T. M. et al. Assessing the overlap between immunisation and other essential health interventions in 92 low- and middle-income countries using household surveys: opportunities for expanding immunisation and primary health care. eClinicalMedicine 42, 101196 (2021).

9. Galles, N. C. et al. Measuring routine childhood vaccination coverage in 204 countries and territories, 1980–2019: a systematic analysis for the Global Burden of Disease Study 2020, Release 1. Lancet 398, 503–521 (2021).

10. Figueiredo, A. de et al. Forecasted trends in vaccination coverage and correlations with socioeconomic factors: a global time-series analysis over 30 years. Lancet Glob. Heal. 4, e726– e735 (2016).

11. UNICEF and WHO. Progress and Challenges with Achieving Universal Immunization Coverage, UNICEF and the World Health Organization, July 2020. https://www.who.int/immunization/monitoring_surveillance/who-immuniz.pdf (2019).

12. Council on Foreign Relations. The U.S. War in Afghanistan. https://www.cfr.org/timeline/us-war-afghanistan.

13. United Nations Office for the Coordination of Humanitarian Affairs. Rohingya Refugee Crisis | OCHA. https://www.unocha.org/rohingya-refugee-crisis.

14. The DHS Program - login_main. https://dhsprogram.com/data/dataset_admin/login_main.cfm?CFID=28539468&CFTOKEN=f150420d72cebd9c-1577BB01-F01A-0CAE-BED1E8C2D336A42A.

15. Schneider, A., Hommel, G. & Blettner, M. Linear Regression Analysis. Dtsch. Arztebl. 107, 776– 782 (2010).

16. Akoglu, H. User’s guide to correlation coefficients. Turkish J. Emerg. Med. 18, 91–93 (2018).

17. Schober, P. & Schwarte, L. A. Correlation coefficients: Appropriate use and interpretation. Anesth. Analg. 126, 1763–1768 (2018).

18. Jost, S. LinearCorrelation. https://condor.depaul.edu/sjost/it223/documents/correlation.htm.

