## Supplemental Vaccination Results for "Assessing the Relationship Between Zero-Dose Communities and Access to Selected Primary Healthcare Services for Children and Pregnant Women in Emergency Settings"

Vaccination Result Supplemental

It was observed for nearly all cases in DRC that the linear association was stronger for all vaccines examined when mothers' responses were included in the estimation of vaccination coverage (*All Sources* data) compared against the relationship observed when only using coverage information if it came from a vaccine card (Card Only data). The notable exception to this is in the case of Polio3 where at the national level the card only data produced a fit with an $R^2$ value larger than the *All Sources* data. Furthermore, it was observed that the relationship was at least as strong and often stronger in the specific subnational regions where conflict is more apparent compared to the relationship seen throughout all subnational regions in the country. These trends for MCV are the same observed for DTP1. [Figure S1]. Likewise, in Afghanistan a stronger linear association was observed when examining the *All Sources* data compared against the vaccination *Card Only* data. Unlike with the DTP1 relationship, here this trend was observed for all cases [Figure S2]. As with the survey results from both DRC and Afghanistan a stronger linear association was observed when examining the *All Sources* data compared against the vaccination *Card Only* data for MCV similar to that seen for DTP1 in Bangladesh. In this case all $R^2$ values were increased. Especially noticeable were the individual relationships between DTP1, Polio1 and Polio3. For these three vaccines, the $R^2$ values were below the cut off for a weak linear relationship to be observed when only vaccination card data was considered, but after the inclusion of responses from mothers all displayed linear relationships. We note here that while BCG did not meet the weak linear association cut off metric, the inclusion of the mothers' response still resulted in nearly a fourfold increase in $R^2$ value.

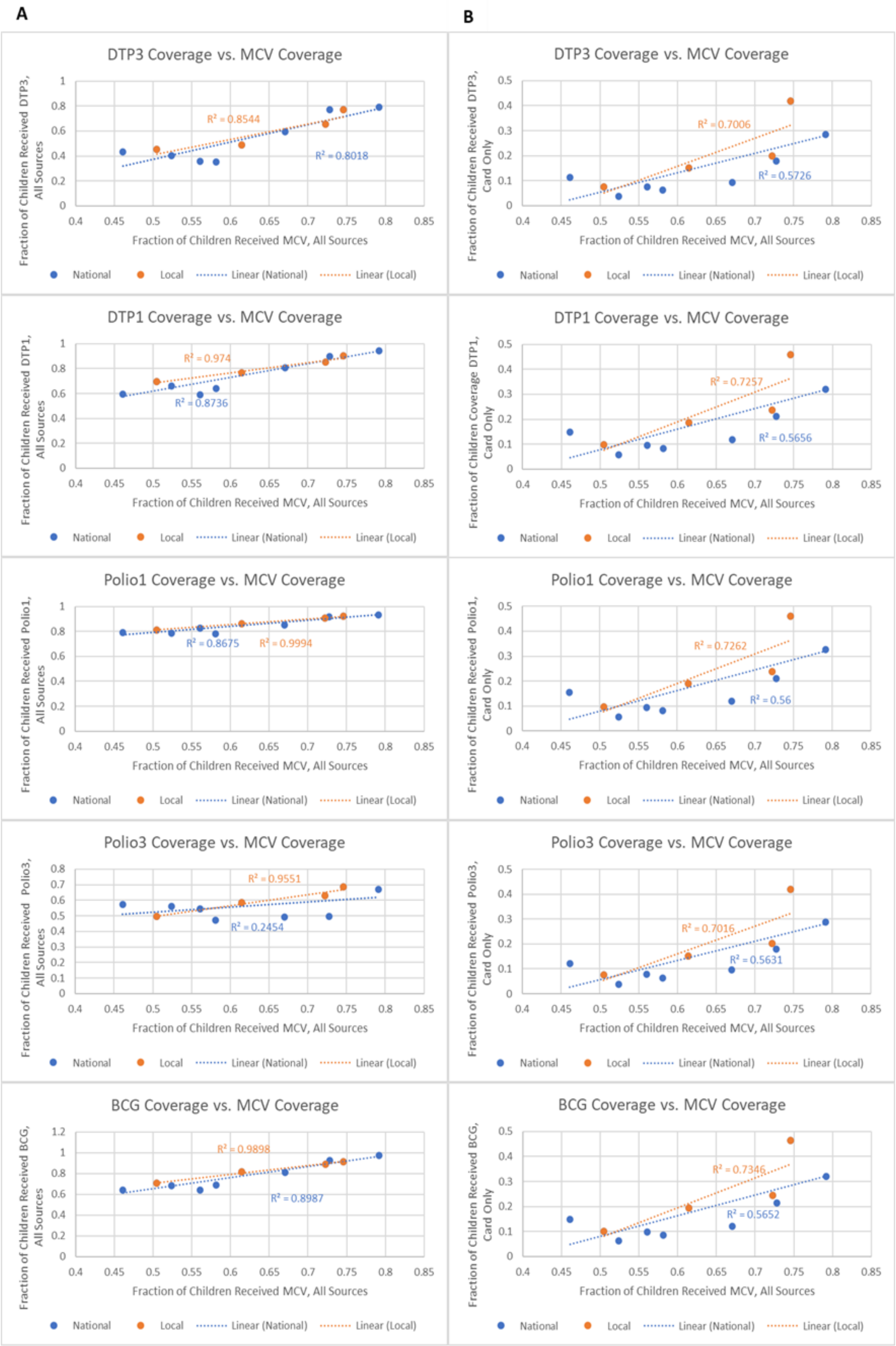


**Figure S1. Linear regression between MCV vaccination coverage e and coverage for other common vaccines in DRC**. Column A shows how the inclusion of mothers' responses improve the linear fit compared to the relationships in column B which only considered a child vaccinated if their vaccination card contained the appropriate vaccine information. In this case, a difference was observed between the strength of the relationship observed in regions with conflict compared to the entire country.

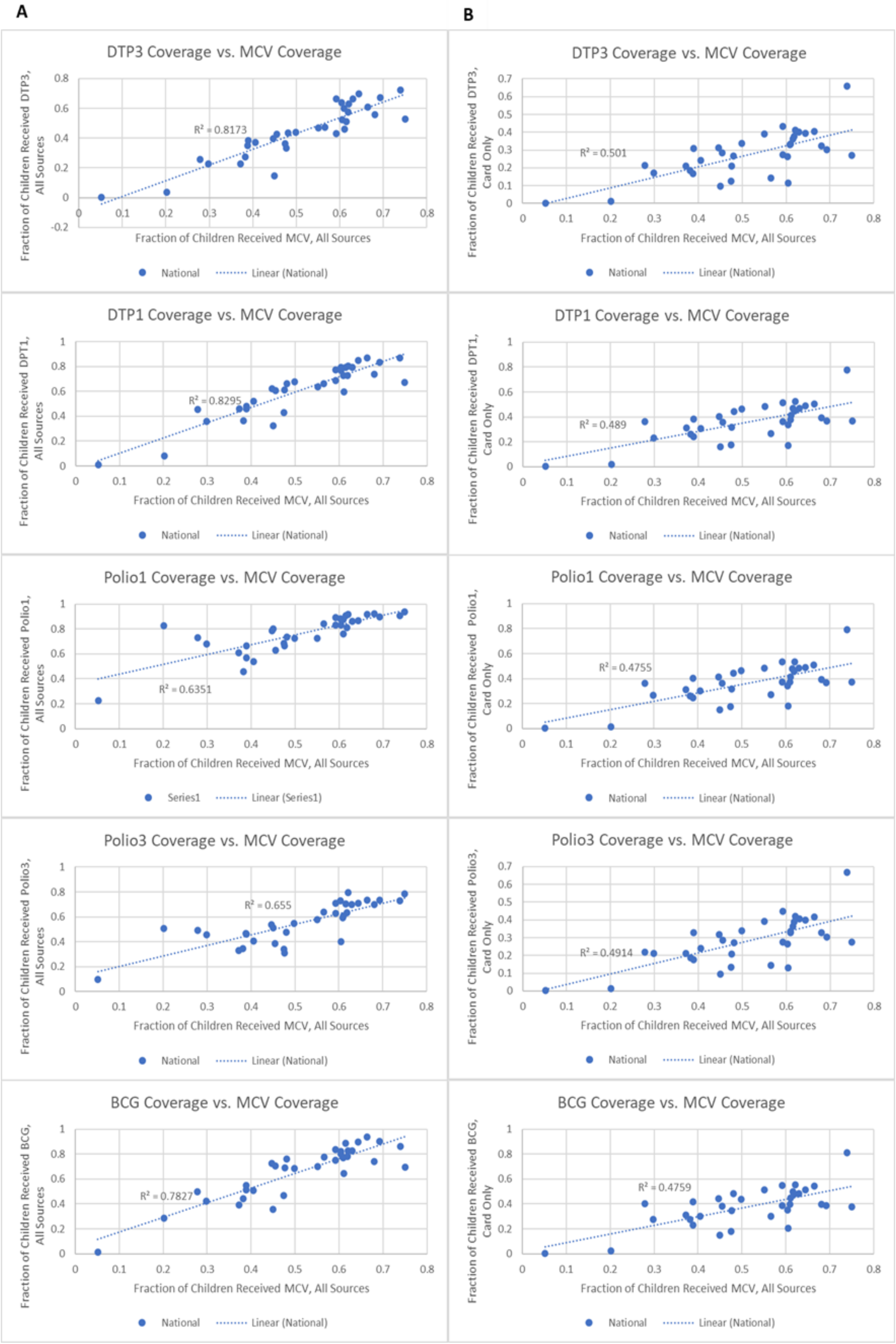


**Figure S2. Univariate linear regression between MCV vaccination coverage and coverage for other common vaccines in Afghanistan.** Column A shows how the inclusion of mothers' responses improve the linear fit compared to the relationships in column B which only considered a child vaccinated if their vaccination card contained the appropriate vaccine information. Unlike for DTP1 This trend did not breakdown for either Polio vaccine dose.

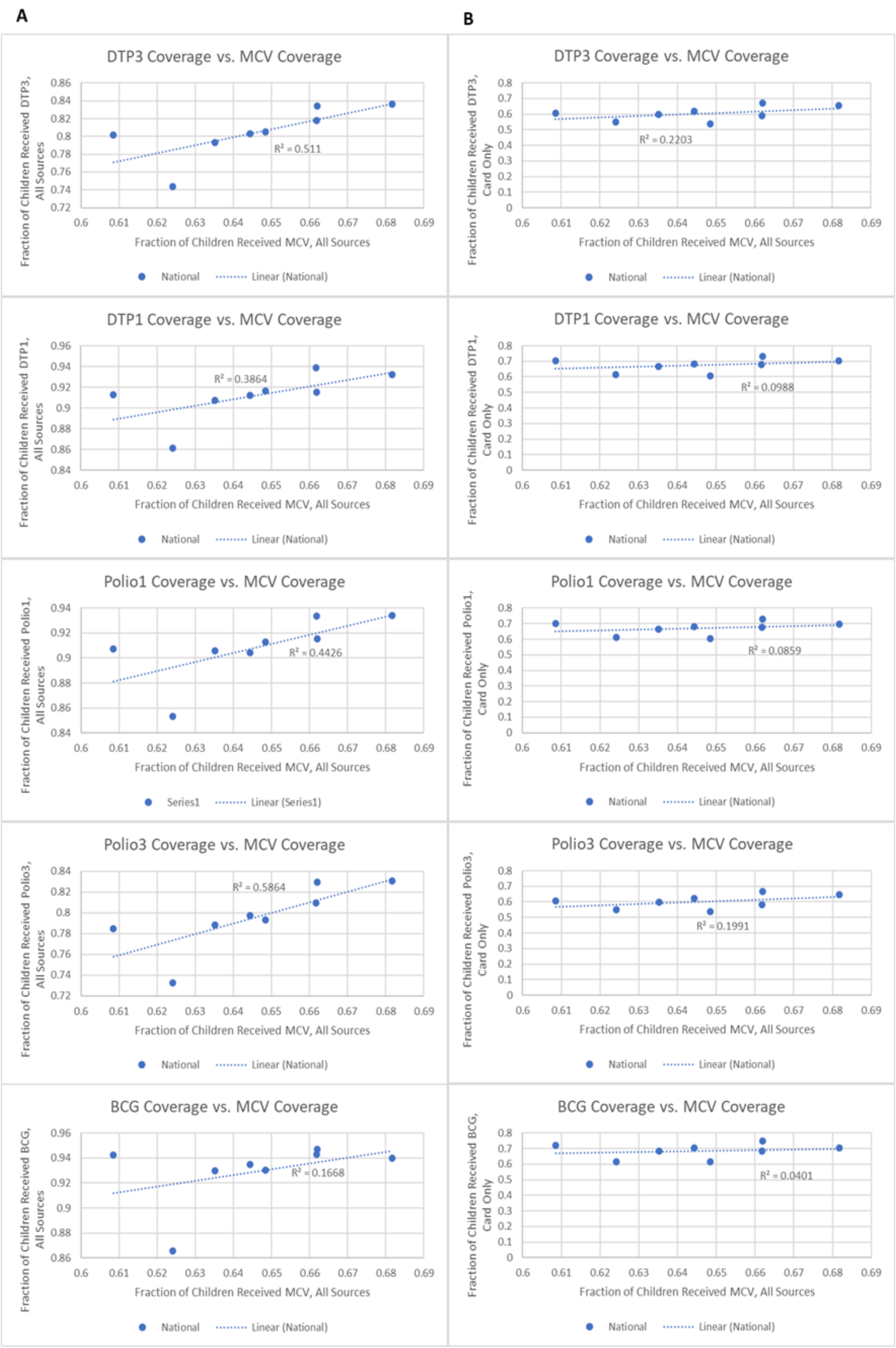


**Figure S3. Univariate linear regression between MCV vaccination coverage and coverage for other common vaccines in Bangladesh.** Column A shows how the inclusion of mothers' responses improved the linear fit compared to the relationships in column B which only viewed a child as vaccinated if their vaccination card contained the appropriate information. This trend was especially apparent for DTP1, Polio1 and Polio3 as the card only metric led to no linear relationship being observed, while the *all sources* data saw linear associations for all of these vaccines.
